# Adherence Behaviors to Prevent COVID: The Role of Anxiety and Prosocial Behaviors

**DOI:** 10.1101/2023.09.18.23295630

**Authors:** Silvia Corbera, Amanda M. Marín-Chollom

**Author notes:** **Corresponding Author:** Silvia Corbera, PhD, Associate Professor, Department of Psychological Sciences, Central Connecticut State University, Marcus White, Room 207, 1615 Stanley Street, P.O. Box 4010, New Britain, CT 06053.

## Abstract

**Objective:** In situations of acute stress, individuals may engage in prosocial behaviors or alternatively, individuals may engage in risk taking self-oriented behaviors. The COVID-19 pandemic created large stress-promoting conditions that impacted individuals’ decisions to adhere to COVID-19 preventative behaviors. The aim of this study is to examine the relationship between anxiety during the pandemic and adherence behaviors to prevent the spread of COVID-19, and the moderating influence prosocial behaviors.

**Method:** 54 undergraduate students completed online questionnaires during the second wave of the pandemic: prosocial behaviors, anxiety, and COVID-19 preventive behaviors. Moderation analyses were conducted using Process in SPSS.

**Results:** Results demonstrated a statistically significant interaction of public prosocial behavior with state anxiety (β = -.17, p=.01) predicting engagement in COVID-19 preventative behaviors. At low levels of anxiety, low levels of prosocial public behaviors were associated with higher engagement in COVID-19 preventative behaviors. In contrast, high levels of public prosocial behavior were associated with lower engagement in COVID-19 preventative behaviors at low levels of anxiety.

**Conclusion:** Results provide information that can aid at in the creation of anxiety reducing interventions that could increase adherence to COVID-19 preventative behaviors.

## Introduction

The understanding of human behavior under stress and in situations of anxiety has been an extensive topic of research (Starcke & Brand, 2012; Nowacki et al., 2018). Ample evidence suggests that acute stress affects general cognition, executive functioning, working memory (Shields et al., 2016; Wolf et al., 2015; Duesenberg et al., 2016); social-emotional information processing and social and prosocial behavior (Frisch et al., 2015; von Dawans et al., 2021; Gonzalez-Liencres et al. 2016). Stress also affects decision-making skills (Starcke & Brand, 2012, 2016; Nowacki et al., 2018, de Visser et al., 2010; Soshi et al., 2019); however, this relationship depends on several factors, such as the type of decision-making situations (Starcke & Brand et al., 2016), time pressure (Soshi et al., 2019) and the individual characteristics of the participants, such as gender (van de Bos, 2009; Nowacki et al., 2018).

Evidence has also shown that social behavior and social decision making can be influenced by stress, however, studies are conflicted in this area. Some studies have shown that in situations of acute stress individuals might engage in more pro-social and empathetic behaviors such as pro-social trustworthiness and sharing (von Dawans, 2012, 2019, 2021; Wolf et al., 2015); however, on the other hand, other studies suggest that in situations of stress, individuals might engage in unfavorable and risk taking self-oriented behaviors and distrust (Bendahan et al., 2017; Steinbeis et al., 2015). Most of the research on the effects of stress in decision-making has been conducted in laboratory studies using stress-inducing paradigms, in which the stress induced was either physical (e.g., cold pressure test in which participants immerse their hand in a bucket of cold water) or psychosocial (e.g., Trier Social Stress Test (TSST) in which participants perform a mock job interview). However, to our knowledge, just one study has investigated decision-making in in the context of the COVID-19 pandemic as the stressor (Romero-Rivas & Rodriguez-Cuadrado, 2021). Romero-Rivas and Rodriguez-Cuadrado, 2021, examined whether the psychological impact generated by the COVID-19 pandemic influenced decision-making processes by presenting participants with four decision-making tasks (the dictator game, framing problems, utilitarian/deontological moral dilemmas and altruistic/egoistic moral dilemmas). This study showed that higher levels of psychological impact were related with safer responses in framing problems and with more deontological/altruistic responses to moral dilemmas (Romero-Rivas & Rodriguez-Cuadrado, 2021). Therefore, suggesting that the psychological impact of COVID-19 affected decision-making processes by participants showing safer and more altruistic responses using laboratory decision-making tasks. Given the influence of stress in decision-making process (Starcke & Brand, 2012; Nowacki et al., 2018) it is imperative to understand this relationship further, especially during the COVID-19 pandemic as it can aid understanding the factors that influence compliance and adherence to behaviors to prevent the spread of COVID-19 and its variants.

The worldwide COVID-19 pandemic placed humanity in a devastating global health challenge that required the population that adapt to a rapid changing situation. Gruber and colleagues (2021) conceptualized the COVID-19 pandemic as a multidimensional complex stressor that affected both the individual and family and multiple societal layers, with toxic social stressors, such as social isolation and financial loss (Gruber et al., 2021). Stress research has shown, that situations that are uncontrollable, uncertain, and with social evaluative threat elicit high stress responses (Dickerson et al., 2004). The COVID-19 pandemic created large stress-promoting conditions, such as uncertainty, life-threat, loss, and long exposure to anxiety-inducing information (Gruber et al., 2021) generating extensive detrimental psychological and mental health consequences (Gruber et al., 2021; Luo et al., 2020). A recent meta-analysis by Luo and colleagues (2020) reported a prevalence of anxiety of 33% and depression of 28% of the population during the COVID-19 pandemic and a study by Ebrahimi and colleagues (2021) reported that the prevalence of clinical anxiety to increase 2 to 3 times during the COVID-19 pandemic in comparison to estimations with other similar samples before the pandemic. In addition, the COVID-19 pandemic has increased the prevalence of anxiety amongst undergraduate college students (Rudenstine et al., 2020; Lee et al., 2021) specially amongst the lower socio-economic groups (Rudenstine et al., 2020).

To prevent the spread of the coronavirus, public health measures were introduced that involved the engagement in new behaviors and the population was expected to adhere to these preventive behaviors (e.g., limiting social distance; the use of masks). Decisions made under different levels of risk and ambiguity can be affected differently by stress (Starcke & Brand et al., 2016; Brand et al.,2006) and during the stress-promoting conditions inherent in the COVID-19 pandemic, the population was confronted with making high-risk decisions about their behaviors, especially on decisions related to preventing the spread of COVID-19.

A few studies have examined the predictors of adherence to preventive behaviors to spread the coronavirus (Pollak et al., 2020; Ebrahimi et al., 2021; Pfattheicher et al., 2020; Syropoulos & Markowit, 2020). Recent research by Pollak and colleagues (2020) showed that predictors for non-adherence were high levels of current distress, and by risk factors such as male gender, not having children, ADHD symptoms, smoking and high levels of risk-taking behavior. Similarly, Ebrahimi and colleagues (2021) reported that female gender, older age, worry about significant others, mandatory adherence and altruistic attitude were associated with higher adherence and current employment was associated with lower adherence. Besides altruistic attitudes, empathy, fairness and gratitude have also been associated with higher levels of adherence to COVID-19 preventative behaviors (Ebrahimi et al., 2021; Pfattheicher et al., 2020; Syropoulos & Markowit, 2020).

To our knowledge no studies have examined the relationship between anxiety and adherence to behaviors to prevent the spread of COVID-19 with the moderating role of prosocial tendencies. Therefore, the present study aimed to investigate 1) the relationship between anxiety during the COVID-19 pandemic and adherence behaviors to prevent the spread of COVID-19, and 2) the moderating influence of prosocial tendencies on this association.

It is hypothesized that individuals with a high anxiety during COVID-19 pandemic are more likely to adhere to the behaviors to prevent the spread of COVID, than individuals with low anxiety and this relationship will be stronger for individuals with high prosocial behavior tendencies than those with low prosocial behavior tendencies.

## Methods

This study was IRB approved and online passive consent was obtained from participants. Participants were undergraduate students recruited at a university in the Northeastern United States through the Psychology Department online subject pool that is managed by SONA. Students from all different majors taking psychology courses were eligible to sign-up to participate in this study and received credit in their courses for participation. Participants were granted 1 SONA credit for 15-30 minutes of their time to complete the questionnaire administered online via Select Survey. Data was collected during the Spring 2021 semester of the COVID-19 pandemic from February 2021 to May 2021. The data reported in this study is a subset of the measures collected in this larger study conducted during those months. In addition to the measures described below, basic demographics of age, race/ethnicity, class standing, gender, and employment status were also collected

### Measures

#### Prosocial Behaviors

The 23-item Prosocial Tendencies Measure (Carlo & Randall, 2002; Carlo et al., 2002) was used to measure six types of prosocial behaviors: compliant (2-items), dire (3-items), altruistic (5-items) public (4-items), emotional (4-items) and anonymous (5-items). Compliant is when prosocial behaviors are done because others asked for help (e.g., “I never hesitate to help others when they ask for it.”). Dire is when prosocial are done to help others under emergency or crisis situations (e.g., “I tend to help people who hurt themselves badly”). Altruistic is helping others and expecting little to nothing in return (e.g., “I often help even if I don’t think I will get anything out of helping.”). Public is when prosocial behaviors are completed because others are watching with the motivation to receive external approval (e.g., “Helping others when I am in the spotlight is when I work best). Emotional is when individuals help other because they are under a highly emotional state (e.g., It is most fulfilling to me when I can comfort someone who is very distressed). Anonymous is when prosocial social behaviors are enacted intentionally without other people’s knowledge (e.g., I tend to help needy others most when they do not know who helped them). Items were answered on a 5-point Likert Scale from 1-*does not describe me at all* to 5-*describes me greatly.* Six total sum subscales were calculated and used in analysis. Higher scores indicate greater prosocial behavior for each subscale.

#### Anxiety

The State-Trait Anxiety Inventory (STAI) was used to measure both trait and state anxiety (Spielberger et al.,1983). The 40-item inventory uses 20 items for trait anxiety (e.g., “I am a steady person”) and 20 for state anxiety (e.g., “I feel at ease””). From the 40 items, 19 are reversed scored. All items in this study were rated on a 4-point Likert from *not at all* to *very much so*. Two total sum subscales were calculated, and higher scores indicate greater anxiety for both subscales.

#### COVID-19 Preventative Behaviors

The COVID-19 International Survey (CIS) from the PhenX Toolkit (2020) was used to collect data on what preventative COVID-19 behaviors participants were engaging in. The 23-items within the survey specific to what frequency individuals are engaging in COVID-19 preventative behaviors were used for this study (e.g., hand washing, mask wearing physical distancing, avoiding social gatherings, self-quarantining after travel, self-quarantining if infected or likely infected were used). These items are answered on a 4-point Likert scale from *never* to *most of the time*, with the additional option of “don’t know/I prefer not to answer/Not applicable.” One total sum score was calculated, and higher scores indicated higher engagement in COVID-19 preventative behaviors.

#### Analysis

Bivariate correlations between the main variables of interest were first examined. Then moderation analyses were conducted using Process version 3.5 within SPSS Version 27. Bootstrapping was set to 5,000 and confidence interval to 95%. Mean deviated predictor variables (i.e., prosocial behaviors and anxiety types) were created and used for all moderation analyses. A total of 12 separate models were tested. One for each type of prosocial behavior (6) interacting with the two subtypes of anxiety measured predicting engagement in COVID related preventative behaviors. All significant interactions were plotted, using 1 standard deviation and above the mean for prosocial behaviors, for interpretation reasons.

## Results

Participants were (n=54) college students (M=20.74, SD=5.16) and mostly enrolled full-time at the university (n=43), employed (n= 43), and living at home with their parent, relative or guardian (n=40). Most of the sample self-identified as women (n=43; men =9; Transgender=1), and white (n=34; Hispanic/Latino/a=11, Black/African American=7, and Asian =2).

Participants on average reported high levels of anxiety symptoms on both the trait (M= 51.05; SD=10.37) and state subscales (M= 50.63; SD=11.11). Using the recommended cut-off of 40 to determine clinically meaningful anxiety symptomology, most of the sample either met or exceeded the criteria (state anxiety 87% and trait anxiety 86%). On average, participants highly adhered to engage in COVID preventative actions and behaviors (M =60.28;13.29). Over fifty percent of the sample engaged in the following behaviors “most of the time”: wearing a face (90.7%), using hand sanitizer (83.3%), handwashing with soap and water (81.5%), coughing/sneezing into elbow (81.5%), self-quarantining if they have or believe they have the virus (77.8%), self-quarantining if they are returning from a trip (63%), coughing/sneezing into a tissue and throwing it away and washing hands (68.5%), staying 6 feet apart from other people (53.7%), avoiding large social gatherings (53.7%), and avoiding any non-essential travel (53.7%). See Table 1.

**Table 1.** Engagement in COVID-19 Preventative Behaviors in the Past Seven Days (n=54)

| Action or Behavior | % “Most of the Time” |
| --- | --- |
| 1. Wearing a face mask | 90.7% |
| 2. Use Hand Sanitizer | 83.3% |
| 3. Handwashing with soap and water | 81.5% |
| 4. Coughing/sneezing into your elbow | 81.5% |
| 5. Self-quarantining if you have or believe you have the virus | 77.8% |
| 6. Coughing/sneezing into a tissue throwing it away and wash hands | 68.5% |
| 7. Self-quarantining if you are returning from a trip | 63.0% |
| 8. Staying 6 feet apart from other people | 53.7% |
| 9. Avoid large social gatherings | 53.7% |
| 10. Avoiding any non-essential travel | 53.7% |
| 11. Avoiding going out to bars/pubs | 48.1% |
| 12. Avoiding using public transportation | 44.4% |
| 13. Staying/working at home | 38.9% |
| 14. Avoiding playdates | 29.6% |
| 15. Exercise outside alone or with people you live with only | 29.6% |
| 16. Avoiding going to restaurants | 22.2% |
| 17. Avoiding taking your children to the park | 20.4% |
| 18. Avoiding all social gatherings (large and small) | 18.5% |
| 19. Wearing gloves every time you go out of your home | 11.1% |
| 20. Avoiding going to the grocery store or pharmacy | 11.1% |
| 21. Avoiding opening the mail or delivered goods | 11.1% |
| 22. Avoiding getting take-out food or delivery | 9.3% |
| 23. Avoiding going for walks | 2.0% |

None of the prosocial behavior types or anxiety types were associated with the engagement in COVID preventative behaviors. See Table 2 descriptive and correlations of study variables. However, in the one model tested with public prosocial behavior, the interaction of public prosocial behavior with state anxiety predicting engagement in COVID preventative behaviors was statistically significant (β = -.17, *p*=.01), which suggested a cross over effect (See Figure 1). At low levels of prosocial public behaviors, as anxiety increases engagement in COVID preventative behavior increases, in contrast at high levels of prosocial public behaviors as anxiety decreases engagement in COVID preventative behavior increases. In the other 11 models tested none of the interaction effects with prosocial behaviors and anxiety predicting adherence to COVID preventative behavior were statistically significant. (See Tables 3 and 4).

**Figure 1.**
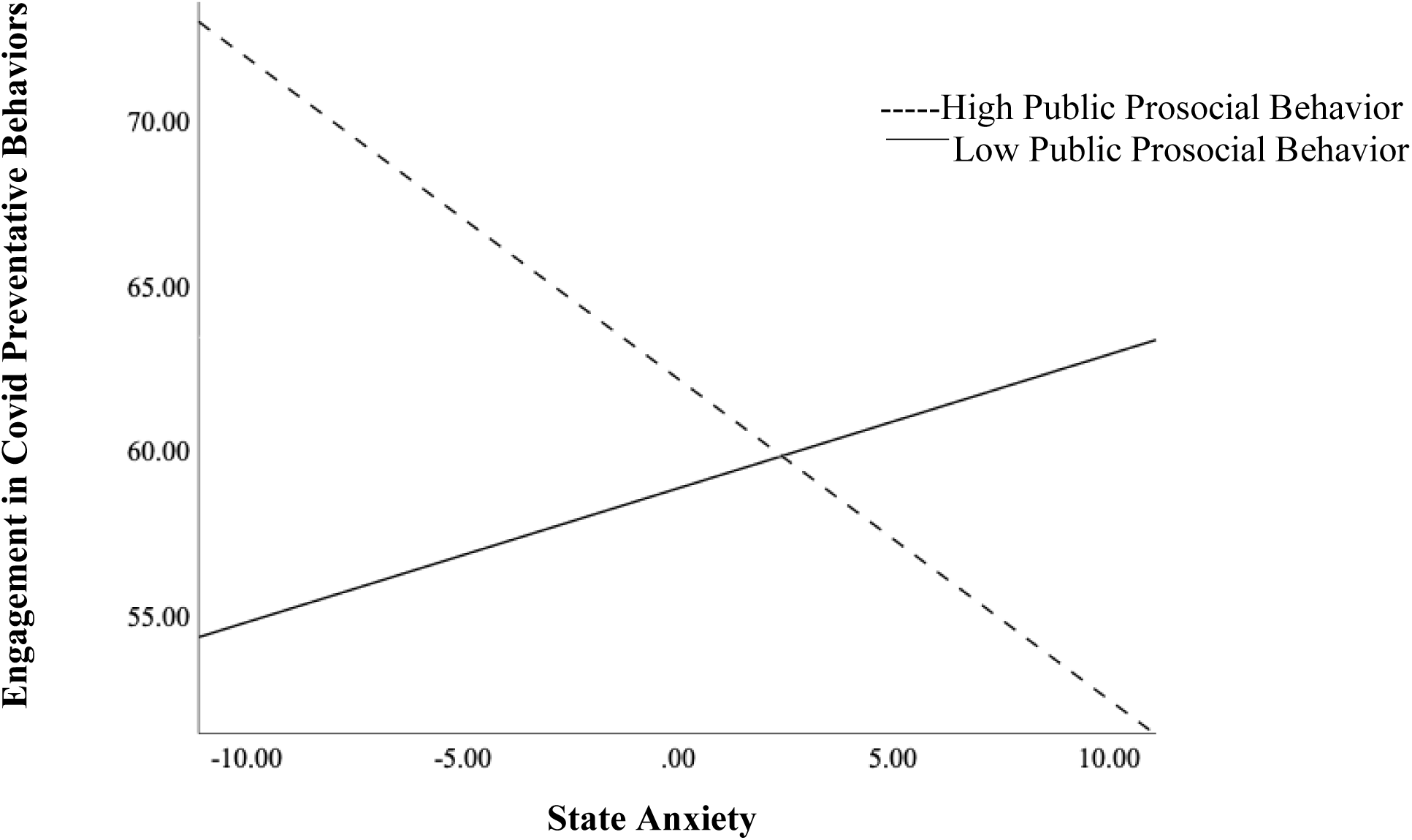
Public Prosocial Behavior Moderating the Association between State Anxiety and Engagement in COVID Preventative Behaviors.

**Table 2.** Descriptive Statistics and Correlations for Study Variables.

| Variable | n | M | SD | 1 | 2 | 3 | 4 | 5 | 6 | 7 | 8 | 9 |
| --- | --- | --- | --- | --- | --- | --- | --- | --- | --- | --- | --- | --- |
| 1.COVID PB | 54 | 60.28 | 13.29 | - | .06 | .01 | -.16 | .04 | -.03 | .16 | -.03 | -.01 |
| 2.Public PT | 54 | 8.26 | 3.48 | - | - | -.11 | -.78** | -.05 | -.26 | .26 | -.03 | .05 |
| 3.Emotional PT | 54 | 13.93 | 3.33 | - | - | - | .09 | .64** | .64** | .49** | -.16 | -.21 |
| 4.Altruistic PT | 54 | 19.83 | 4.15 | - | - | - | - | .02 | .31* | -.20 | -.06 | -.09 |
| 5.Dire PT | 54 | 10.35 | 2.63 | - | - | - | - | - | .64** | .55** | .03 | .11 |
| 6.Compliant PT | 54 | 7.89 | 1.76 | - | - | - | - | - | - | .38** | -.23 | -.20 |
| 7.Anonymous PT | 54 | 14.74 | 4.34 | - | - | - | - | - | - | - | -.13 | -.18 |
| 8.Trait Anxiety | 44 | 51.05 | 10.37 | - | - | - | - | - | - | - | - | .84 |
| 9.State Anxiety | 54 | 50.63 | 11.11 | - | - | - | - | - | - | - | - | - |

*Note.* PB= Preventative Behaviors, PT=Prosocial Tendency, \*\* $p \leq .01$ , \* $p \leq .05$ .

**Table 3.** Association between State Anxiety and Covid Preventative Behaviors Moderated by Six Types of Prosocial Behaviors (n=54)

| Model | Estimate | SE | t | p |
| --- | --- | --- | --- | --- |
| 1. Complaint | -.58 | 1.12 | -.52 | .61 |
| State Anxiety | -.88 | .75 | -1.18 | .25 |
| Complaint X SA | .13 | .11 | 1.19 | .24 |
| 2. Emotional | -.25 | .60 | -.42 | .68 |
| State Anxiety | -.78 | .55 | -1.42 | .16 |
| Emotional X SA | .07 | .05 | 1.49 | .14 |
| 3. Public | .41 | .51 | .80 | .43 |
| State Anxiety | -.33 | .20 | -1.65 | .10 |
| Public X SA | .17 | .06 | -2.60 | .01 |
| 4. Altruistic | -.60 | .44 | -1.38 | .17 |
| State Anxiety | -1.76 | .94 | -1.88 | ..07 |
| Altruistic X SA | -.10 | .05 | 1.88 | .07 |
| 5. Dire | .31 | .73 | .42 | .68 |
| State Anxiety | -.33 | ..48 | -.69 | ..49 |
| Dire X SA | .04 | .06 | .71 | ..50 |
| 6. Anonymous | .51 | .44 | 1.16 | .25 |
| State Anxiety | -.16 | .42 | -.37 | .71 |
| Anonymous X SA | -.02 | .04 | -.48 | .63 |

*Note.* DV= Covid preventative behaviors All variables were mean deviated, except the DV.
SA=State Anxiety

**Table 4.**
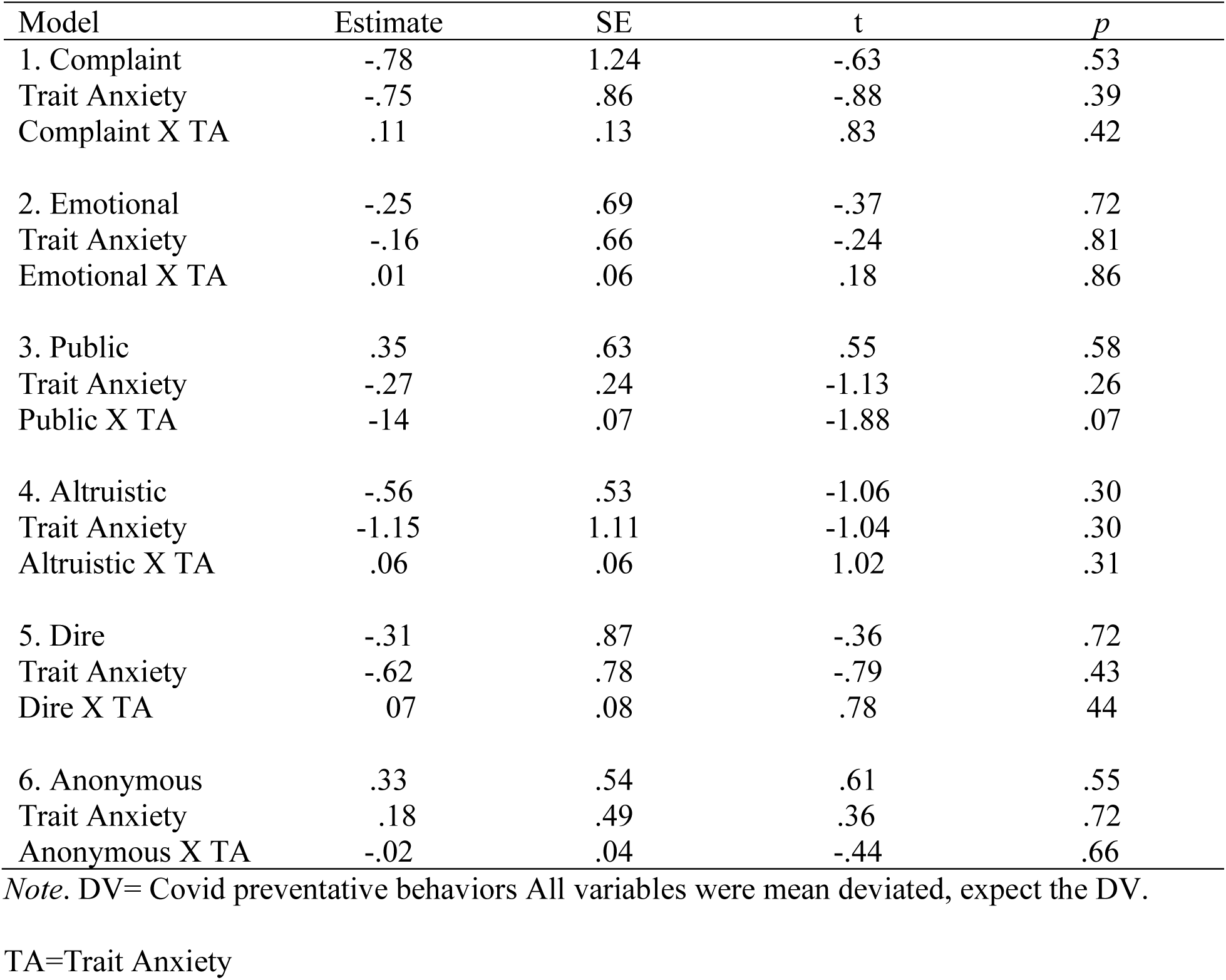
Association between Trait Anxiety and Covid Preventative Behaviors Moderated by Six Types Prosocial Behaviors (n=44)

## Discussion

The present study aimed to examine the association between state and trait anxiety during the COVID-19 pandemic and the adherence behaviors to prevent the spread of COVID-19 (e.g., handwashing, mask wearing), and to investigate the moderating role of prosocial behaviors on this association. Based on previous literature on decision-making under stress promoting conditions (von Dawans, 2012, 2019, 2021; Wolf et al., 2015), we hypothesized that participants with high state and trait anxiety would be more likely to adhere to the behaviors to spread COVID-19 than those with low state and trait anxiety, and that this relationship would be stronger for individuals with high prosocial behavior tendencies, than those with low prosocial tendencies. Because of the exceptional situation created by the COVID-19 pandemic we did not specifically predict that any type of prosocial behavior would show a stronger relationship versus another. Our results revealed that: 1) There were no associations between state and trait anxiety and any of the prosocial behaviors (emotional, altruism, dire, anonymous, public, compliant); 2) Only public prosocial tendencies showed a significant interaction with state anxiety, predicting engagement in COVID-19 preventative behaviors, and suggesting a cross over effect.

Concerning the lack of association between state and trait anxiety and any of the prosocial behaviors, these results were not surprising as we expected that differential levels of prosocial tendencies would relate differently with anxiety when predicting adherence behaviors and therefore that they would operate as a moderator. On the other hand, the interaction that illustrated that public prosocial behaviors moderated the relationship between state anxiety and adherence behaviors to prevent COVID-19 did not support our hypothesis, as the resulting cross-over effect reflected that increased state anxiety related with an increase in adherence behaviors but in individuals with low public prosocial behaviors. However, as shown by the validation study of the PTM of Carlo & Randall (2002), individuals who engage in public prosocial tendencies showed approval seeking behaviors and were oriented towards their own needs (as opposed to other people’s needs). Additionally, public prosocial behaviors were significantly negatively correlated with altruistic prosocial behaviors, which are other-oriented and with sympathy (Carlo & Randall, 2002).

Because our results showed that increased state anxiety, increased adherence behaviors in individuals with low public prosocial behaviors, they illustrated that individual with lower self-oriented tendencies and with less approval seeking tendencies (low public prosocial behavior), with higher state anxiety would show more engagement in adherence behaviors. The opposite pattern was found with individuals with high public prosocial tendencies, in which the increase of state anxiety would decrease their engagement with adherence behaviors. In addition, given the inverse relationship between public and altruistic prosocial behaviors (Carlo & Randall, 2002), these results suggest that individuals low on public prosocial behaviors would show more other oriented behaviors. On the other hand, our study did not find any differential relationship with individuals with altruistic prosocial behaviors and state anxiety and adherence behaviors, and therefore, more research is needed in this area. Our results are in partial agreement with the study of Romero-Rivas and Rodriguez-Cuadrado (2021), which found that high states of psychological impact from the COVID-19 pandemic was related to individuals being more risk-aversed and more altruistic in decision making. Our results showed higher state anxiety was related to more engagement in adherence behaviors to prevent COVID-19 in only the individuals with lower public prosocial behaviors (more other-oriented and less approval seeking behaviors). Therefore, we propose that higher anxiety relates with prosocial decision-making differently depending on the individual characteristics, in which the individuals with more other-oriented/less-approval seeking prosocial tendencies with higher state anxiety would be more likely to adhere more in COVID-19 preventive behaviors. On the other hand, individuals with higher levels of self-oriented/approval seeking behaviors (public prosocial), show less adherence to COVID-19 preventive behaviors with the increase of state anxiety. As the group with high public prosocial tendencies has been reported to be motivated by self-oriented and approval seeking tendencies (Carlo & Randall, 2002), we speculate that situations with high state anxiety would prevent these individuals from engaging in behaviors that would necessitate of other-oriented prosocial behaviors and therefore, would prevent them from engaging in prosocial decision making and in other-oriented behaviors.

Finally, we wanted to highlight that our study sample showed high levels of both state and trait anxiety (87% sample in state anxiety and 86% sample in trait anxiety above 40 cutoff). but only state anxiety related with adherence to behaviors to prevent COVID-19. Given that the study was conducted in the Spring of 2021 of the COVID-19 pandemic, in a college-sample, in such an unprecedented time, it was not surprising that the levels of anxiety were high.

We suggest that more studies are needed to explore the role of each type of prosocial behavior tendencies in addition to a global measure of prosocial tendencies, in the engagement of adherence behaviors to prevent COVID-19. Our results further the current knowledge on the factors that predict the engagement to adherence behaviors to prevent COVID-19 and provide insights on the cultivation and creation of interventions to promote other-oriented vs self-oriented motivations and tendencies and in the creation of anxiety reducing interventions that could increase adherence to COVID-19 preventative behaviors.

## Limitations

Our study has several limitations. First the sampling method used was convenience sampling with undergraduate college population, which could have led to collect data from participants that were more motivated and self-selected. Secondly, the study was conducted online and therefore it was not possible to control for attention and effort in the participants responses. Additionally, the measures were self-report questionnaires that are prone to biases, such as social desirability. Finally, this study was cross-sectional and limits the interpretation of the data.

## Conclusion

To sum up, our results revealed that anxiety affected differently the prosocial-decision making process, as at low levels of prosocial public behaviors, as state anxiety increased, also increased the engagement in adherence in behaviors to prevent the spread of COVID-19. On the other hand, at high levels of prosocial public behaviors, as state anxiety increased, the engagement in adherence in behaviors to prevent the spread of COVID-19 decreased. These results provide valuable information that can aid in the creation of interventions to promote other-oriented motivations and in the reduction of anxiety that could increase adherence to COVID-19 preventative behaviors.

## Contribution Statement

Both authors contributed substantially to each stage of the research process: study design, data collection, data analysis, and drafting and revising the manuscript.

## Data Availability

All data produced in the present study are available upon reasonable request to the authors

